# Combining deliberative engagement with qualitative research to assess patient and caregiver perspectives on curative therapies for sickle cell disease in Africa

**DOI:** 10.1101/2025.07.30.25332426

**Authors:** Daima Bukini, Jennifer Mashaka, Aisha Rifai, Collin Kanza, Kassim Kassim, Rahma Chanzi, Deogratius Maingu, Frederick Luoga, Winfrida Lema, Fatou Konteh, Mohammed Zahir, Elianna Amin, Jonathan Spector, Julie Makani

## Abstract

The vast majority of individuals impacted by sickle cell disease (SCD) live in Africa, where access to potentially curative therapies such as hematopoietic stem cell transplant and gene therapies have been severely limited until now. As part of a broad effort to help prepare scientific, healthcare, and patient communities in Tanzania for the introduction of advanced SCD therapies, we sought to understand perspectives relating to curative options for SCD held by patients and their families. To achieve this, innovative qualitative research methodology was required given the highly variable level of baseline knowledge among patient communities regarding curative treatments for SCD. We therefore adopted an approach of deliberative engagement followed by traditional qualitative methods that facilitated successful identification of participants, development of research tools, and, ultimately, elicitation of valuable insights that enabled in-depth analyses. Our experience demonstrates that, when exploring perspectives relating to complex medical procedures in this population, deliberative engagement substantially enhances the outputs of qualitative research.

## Introduction

The greatest burden of sickle cell disease (SCD) is in Africa where approximately 300,000 babies are born with the disease each year^1,2^. While early diagnosis enables the delivery of comprehensive care treatments that reduce morbidities and mortalities associated with SCD,^3– 5^ until recently the only cure was hematopoietic stem cell transplantation (HSCT) through fully matched sibling or haploidentical donors^6–8^. In late 2023, however, the first two SCD gene therapy products were approved by the United States Food and Drug Authority (FDA), ushering in a new era of promise that cure could be achievable through advanced technological treatments.^9–11^ While the reality is that access to these novel therapies has so far been out of reach for most Africans given cost and logistical challenges, the glimpse of hope for patients and their families in Africa has provided impetus for multi-sectoral stakeholders to prepare the landscape for the introduction of gene therapies on the continent. ^12,13^

Tanzania is estimated to have approximately 11,000 to 16,000 annual births of children born with SCD, ranking fifth in the world for having the highest burden of the disease^14–16^. While there is still limited access to newborn screening and disease modifying therapies such as hydroxyurea, the country has made considerable progress in strengthening comprehensive care services for patients with SCD^17–20^. In 2023, the first HSCT for children with SCD was performed at Benjamin Mkapa Hospital in Dodoma^12^. This progress, further bolstered by the commitment of national health authorities, has helped to drive the interest of patients, families, researchers and clinicians to explore preparedness for gene therapy programs for SCD.

Deliberative engagement (DE) is a participatory approach used across disciplines to facilitate in-depth discussions, decision-making, and research activities^21^. Key features of DE are the provision of information to establish a shared knowledge base among participants that otherwise have varying levels of understanding about a particular topic, and providing the opportunity for discussion and clarification through two-way interactions^22,23^ In healthcare, DE has been extensively utilized to support understanding of complex phenomenon in health sciences which includes facilitating discussions related to the introduction of new medical technologies or treatments. ^24–27^ In Africa, DE has previously been used as an approach to engage with the public on the topic of human genome editing and to explore understanding on genomic research studies. ^2829^ In these cases, DE helped to overcome the challenge that topics were new to participants that had poor baseline familiarity of their practical application to health sciences.

Traditional qualitative research, involving methodology such as In-depth interviews (IDIs) or focus group discussions (FGDs), has led to valuable contributions in the health sciences, with an important facet being the incorporation of patient experiences into clinical research. Indeed, much of what is commonly referred to as empirical bioethics is in fact utilization of qualitative approaches to understand ethical issues in healthcare. A distinction between traditional qualitative research and DE is that the method of inquiry in the former is usually characterized by one-way data collection whereas the latter is designed to allow for more interactive discussions. Traditional qualitative methods have been successfully used to understand the lived experiences of patients with SCD and their families.

Our team has previously utilized traditional qualitative methods to conduct empirical work in the ethics of genetics research and health care ^30,31^. In this study, we incorporate combined DE and traditional qualitative methods in an in-depth investigation of Tanzanian patient and caregiver perspectives relating to curative therapies for SCD. Our experience demonstrates that application of this combined approach enhances methodological rigor when exploring new and complex medical phenomena such as gene therapy programs for SCD. This article describes the process and results relating to the novel methodological approach; the specific findings of the analyses will be the subject of another article.

## Methodology

### Study Setting

The study was implemented by the SCD-Advanced Therapy Program (SCD-ATP) at Muhimbili University of Health and Allied Sciences (MUHAS), in collaboration with the Aga Khan main Hospital (AKH), in Dar es Salaam region in Tanzania. SCD-ATP was established through the sickle cell program at MUHAS in 2021, with the goal of bringing together researchers, clinicians, patients and families to advocate for exploration of advanced therapy interventions for SCD in Tanzania. The scope of SCD-ATP includes exchange transfusion services, Human Leukocyte Antigen (HLA) tests and gene therapy preparedness activities. The Program also initiated the first SCD advanced therapy clinic, which runs once weekly at the Jakaya Kikwete Cardiac Institute (JKCI). A minimum of 5-10 patients are seen weekly and, from May 2022 to December 2023, a total of 383 clinical consultations were conducted at the clinic.

### Study Population

All participants in this study were recruited through a registry of individuals living with SCD that are candidates for advanced therapy. These patients were screened through SCD comprehensive care clinics at Amana Regional Referral Hospital, Temeke Regional Referral Hospital, Mwananyamala Regional Referral Hospital and Muhimbili National Hospital. Screening criteria included: patients with abnormal MRI/ TCD results, previous history of strokes, multiple hospitalizations/blood transfusions, avascular necrosis, and other clinical criteria. Up to December 2023, the registry comprised a total of 157 patients, out of which more than 60% of those registered were involved in this study.

**Table 1.** Demographic characteristics of the study population.

| **Category** | **Number** |
| --- | --- |
| Male | 86 |
| Female | 61 |
| **Total** | **157** |
| Adults > 18 years of age | 55 |
| Pediatrics < 18 years of age | 102 |
| **Total** | **157** |
| Patients on Hydroxyurea | 86% |
| Patients with Insurance | 61% |

### Study Procedures

#### Patient engagement camps

Three patient engagement camps were held between March and June, 2021, at the Aga Khan Hospital. In total, 50 patients were involved in the camp sessions, including adult SCD patients and caregivers for pediatric patients. The camps were an adaptation of deliberative engagement approaches in which open dialogue was facilitated between clinicians, patients and families on topics relating to advanced therapies for SCD and, in particular, HSCT and gene therapies. Since the timing of the camps was concurrent with establishment of exchange transfusion programs for SCD at AKH, the discussions also included exchange transfusion. The camps served multiple purposes. They provided an opportunity for (a) patients and families to obtain a basic understanding of advanced therapies, and (b) exploration to ascertain if they would enable the desired open interactions and engagements between clinicians, researchers, patients, and families.

Each camp had a similar format, consisting of day-long health education modules that reviewed advanced therapeutic options for managing SCD. Initial camp sessions placed emphasis on the manual red cell exchange transfusions offered at the Aga Khan Hospital in the Dar es Salaam region. Presentations by relevant clinical specialists provided comprehensive reviews of laboratory, clinical, nursing and referral processes, in addition to expectations and contraindications. Personnel included individuals from management, clinical, nursing and laboratory departments. Later sessions focused on patient feedback and experiences. Patients were encouraged to share their perspectives, concerns and general feedback on the establishment and prospective uptake of such options to treating SCD within the Tanzanian context. Patient representatives in attendance also had the opportunity to recount their experiences living with SCD. Stakeholders and partners (i.e., Ministry of Health, Tanzania Sickle Cell Alliance, SPARCO Tanzania, SPARCO Clinical Coordinating Center) were also in attendance who reasserted their support and continued involvement in efforts to address SCD.

As part of the camps, patients and caregivers were given a physical tour of hospital facilities (AKH) that a patient with SCD would be likely to visit. Patients were able to visit the radiology, phlebotomy, outpatient, and exchange transfusion units and witness firsthand the services offered on site.

Following the tours, patients and caregivers were organized into groups and asked to share their perspectives on the engagement approach through informal discussions in the absence of researchers and clinicians. Box 1 lists the broad topics that were explored during these discussions with patients and caregivers.

##### Box 1

**Examples of discussion guide explored during the camp sessions**

**Box 1. Examples of discussion guide explored during the camp sessions**
|  |
| --- |
| What do you think worked well today during the discussions with researchers and clinicians? |
| How do you think the discussions were helpful in building your understanding of advanced therapies treatments for SCD? |
| Did you have any previous knowledge about the treatments discussed today? If yes, where from? |
| What are your thoughts on the advanced options for treating and managing SCD discussed today? |
| Are there any concerns or worries you have in relation to these advanced options for treating and managing SCD? |

#### Patient focused advanced therapy workshops

Two workshops were held in June, 2022, and April, 2023, involving a total of 102 patients, caregivers and extended family members. This second series of workshops had a specific focus on furthering participants’ understanding of HSCT and gene therapies. Learnings obtained through the patient camps were used to directly inform the discussion topics at the workshops and the format for facilitating engagements and interactions. The workshops started with a general overview of the SCD-ACT followed by a 30-minute presentation in swahili focused on HSCT for SCD patients, a 30-minute presentation on gene therapy programs for SCD, and a question & answer session. After a late-morning break, the particpants were divided into 5 groups of 8-10 people, each of which was was asked to discuss main themes identified during the patient camps; these included: (a) cost of treatments; (b) access and eligibility; and (c) pre- and post-transplant care and family support required; and (4) concerns regarding potential risks of therapy. The group discussions were moderated by clinicians and researchers who were part of the SCD-ATP. After the group discussions, participants had a one-hour break and then took part in feedback sessions in which a representative from each group gave a summary of the discussions.

### Qualitative approaches: In-depth interviews and focus group discussions

The IDIs and FDGs for the qualitative study components started in July 2023, nearly two years after the first camp session. The extended timeframe provided the research team with sufficient opportunity to support knowledge transfer and awareness for patients and families to enable meaningful engagement for IDIs and FSGs. Moreover, analysis of the data obtained through the camps and workshops was used to develop the qualitative tools that guided the IDIs and FGDs. Participants included in this aspect of the study were purposively selected through the workshops. In total, 81 participants were included in IDIs and FGDs. Eight FGDs involving a total of 55 participants were conducted, four with patients and four with caregivers. The FGDs were composed of 5-8 participants per session. Twenty-six IDIs were conducted with patients (13) and caregivers (13). Out of the 81 participants in IDIs and FSGs, 27 particpants were male (21 patients, 6 caregivers) and 54 participants were female (20 patients, 34 caregivers). Table 2 lists demographic descriptions of participants.

**Table 2.** Demographic characteristics of the participants included in the qualitative study.

| Variable | Category | n (%) |
| --- | --- | --- |

|  |  |  |
| --- | --- | --- |
| **Gender** | Male | 27 (33.3) |
|  | Female | 54 (66.6) |
| **Participants** | Caregivers | 40 |
|  | Patients | 41 |
| **Age** | 10-20 | 12 (14.8) |
|  | 21-30 | 22 (27.2) |
|  | 31-40 | 26 (32) |
|  | 41-50 | 17 (20.9) |
|  | 51-60 | 3 (3.7) |
|  | 61-70 | 1 (1.2) |
| **Occupation** | Small business | 40 (49.3) |
|  | Formal employment | 1 (1.2) |
|  | Full time mothers | 18 (22.2) |
|  | Retired | 7 (8.6) |
|  | Students | 6 (7.4) |
|  | Did not identify | 9 (11.1) |
| **Level of education** | Primary | 34 (41.9) |
|  | secondary | 30 (37.03) |
|  | High school | 1 (1.2) |
|  | College | 3 (3.7) |
|  | University | 5 (6.2) |
|  | Did not identify | 9 (11.1) |
| **Number of children with SCD (caregivers)** | One child | 22 (27) |
|  | Two children | 11 (13.5) |
|  | Three children | 4 (4.9) |
|  | Four children | 5 (6.2) |
|  | Did not identify | 5 (6.2) |

#### Box 2

**Examples of the FGD and IDI topics utilized during the qualitative study? Understanding, perceptions and general views**

Box 2. Examples of the FGD and IDI topics utilized during the qualitative study
|  |
| --- |
| Understanding, perceptions and general views |
| • What are your thoughts on the curative therapies for sickle cell disease including gene therapies and stem cell transplantation<br>• Can you briefly explain your understanding of the therapies? |
| **Sources of information** |
| • What information do you believe is important to know regarding each one of these therapies? Exchange transfusion/ stem cell transplant/ gene therapy<br>• What did you think initially about gene therapy, and how did these ideas change over time?<br>• Tell me what you have heard from different people about gene therapy. Given what you've heard, what do you think about it? What do you think you'd do? |
| **Decision-making, preparation and information** |
| • If the gene therapy was available for you or your child, how would the decision-making process be the same or different?<br>• What do you think you need to prepare for any one of these services?<br>• What factors would most influence your use of either gene therapies and/or stem cell transplantation as a mode of treatment? For example: cost, time, location, tradition, culture and religion. If it were up to you, which between the two would you choose, and why?<br>• BMT requires a caregiver to be in the hospital for 3 months and come monthly for 6 extra months for follow up, tell us how the family will negotiate the different roles and obligations in the households and hospitals. Specifically, please share your perspective on caregiving roles at homes and hospitals? How financial obligations will be managed in the families, food, school fees? How are men and women impacted differently during the decision making? |
| **Worries and concerns** |
| • What problems do you believe could arise in the process of using any one of these treatment options?<br>• How do you think they can be solved?<br>• Do you anticipate/foresee any ethical issues in relation to these therapies?<br>• What are your thoughts about the risks of treatment? |

### Data management and analysis

Data were analyzed through thematic content analysis. Except for the patient camp sessions, all sessions were audio recorded and transcripts were analyzed by researchers that were directly involved with the project (DB, CK, KK, RC, AR). Each researcher studied the transcripts and assigned a unique code to every paragraph relating to interviews or discussion. The research team then met in person regularly on a weekly basis to discuss the transcripts and coding exercises. Research team members compared the outputs of their coding approach discussed with each other why each code had been applied. One researcher (KK) was then responsible for compiling all code descriptions and related quotes in a table for the purpose of developing a code book. The codebook was reviewed by the team, and after considerable review and inputs, it was used to code all subsequent transcripts.

## Results

Combining the DE approach with IDIs and FGDs was found to substantially enhance the study conduct, the perspectives provided by patients and caregivers, and the final analyses. Details of observed benefits of this methodology are described below.

### Deliberative engagement greatly facilitated the identification of participants for IDIs and FGDs

Potential participants for the IDIs and FGDs were initially identified during the camp and workshop sessions, and each of the 81 participants in the IDIs and FGDs attended at least one of the DE sessions held during the course of the study. Purposive sampling criteria used to identify participants included age of the participants to ensure young adults were included, given that 70% of the study population was children and it was desired to ensure young adult patients with SCD were adequately represented. Most of the caregivers that attended the camp sessions were mothers of children with SCD, which was expected given that mothers typically assume the primary caregiving role in families in the study’s catchment area.^32,33^ The researchers intentionally approached several fathers (i.e., male caregivers) of children with SCD that attended the DEs to participate in IDIs or FGDs, which resulted in a signficantly larger proportion of male caregiver participants than would have participated in the absence of the DE approach.

### Informing the development of the qualitative tools: IDI and FGD guide

The DE sessions facilitated development of qualitative tools used to guide IDIs and FGDs. The insightful perspectives received from participants during the camps and workshops enabled the research team to understand key questions that were of particular interest to patient and caregivers regarding advanced therapy interventions for SCD with a focus on gene therapy programs. The preliminary engagements also enabled the research team to understand how best to frame questions and what specific questions were most relevant to each group of stakeholders.

The examples below illustrate insights from FGD participants that helped to inform the set of questions related informed consent for advanced therapies.

> *“Ah, you were not clear at first, when you say what are you thinking about advanced therapies, it is not clear, the question should be specific on bone marrow transplant and gene therapy.” (Pre-test FGD_Patients_12 April 2023)*

> *Aha, now we understand you. That’s why when you ask how to understand the components of an informed consent form, I think we need to discuss and ask, ‘What do you think about these questions?’ You have to use very soft language because there are mothers who only went up to fourth grade and they have children with sickle cell and others are sick, So there are things that if we don’t simplify, they will see it as if we are talking about new things from another world. At least we try to make it easier for others, so we need to look at making it easy and friendly. Sometimes people get tired of seeing so many difficult questions.” (Pre-test FGD_Patients_12 April 2023)*

### Guiding the development of proper Kiswahili translation

Kiswahili, also known as Swahili, is the official national language in Tanzania and is used by a majority of the population. During the camp and the workshop sessions, study researchers had a specific interest in learning from patients and caregivers about simple Kiswahili terms that could be used to explain unfamiliar scientific terms (e.g., “*DNA*,” “*genes*,” etc.) that are frequently referred to when educating about advanced therapies for SCD. Based on inputs received from participants, the research team was able to construct a list of simple Kiswahili terms that were then used during the IDIs and FGDs to facilitate understanding of relatively sophisticated concepts. For example, the recognized Kiswahili translation for the word *“genes”* is “*vinasaba”.* However, the word “vinasaba” is not commonly used in Kiswahili language and therefore majority of the participants did not understand its meaning. Through the DEs, participants suggested the Kiswahili word *“chembechembe”* would actually be helpful to use in this context as as adjunct to “*vinasaba”* (the usual translation of *“chembechembe”* to English is “*particles.”*)

The example below illustrates how a participant attempted to understand the meaning of *“bone marrow”* in terms that were more familiar and simple to them.

> *“Thank you. Blood is produced inside the bones, known in English as the bone marrow. I’m not sure of the Swahili term, but to explain it, I would say, let’s take a chicken bone at the end joint. Inside the joint, there’s something like a sponge that you can’t swallow if you chew it; it resembles spinach with red, white, and a bit of black inside the sponge. That’s the marrow. That’s how I understand it. (FGD 1_Patients_27 Jul 2023)*

### Progressive increase in knowledge that supported enhanced participant reflections

From the camps and workshops to the IDIs and FGDs, a steady increase in awareness and understanding of new therapies was observed among study participants. For example, as one participant related:

> *“We were called to a seminar where we were informed about the treatment involving transplantation and blood transfusion. And I was able to understand based on the explanation about bone marrow transplant it is well understood” (IDI 7_Patient_12 Oct 2023)*

This progress was also evident based on the types of questions asked by paticipants and the depth of discussion during FGDs, indicating a greater comprehension of topics than was observed in the initial sessions. For example, during the first camp sessions when the intended focus of discussions was introducing the concept of advanced therapies, common questions asked by patients were rather focused on current treatments received at health centers. Families were interested to better understand causes of delays in currently available routine health services:

> *“I had to come all the way to the hospital. How many hours are those? And still I had not received any treatment yet.” (FGD 4_Patients_16 Jan 2024)*

However, by the time of the workshops, the focus of questions asked by patients and began to shift to advanced therapy programs. For example, participants asked about eligiblity criteria to participate in gene therapy programs (i.e., how would it be decided which patients receive the treatment first). This was exemplified by questioning from an older patient that was concerned about potential age limitations for both HSCT and gene therapy:

> *“… Even us (older patients) want to live.. Now the stem cells, I agree, the children should get. But am I not allowed to receive exchange blood transfusions or gene therapies?” (Arx Patients Focused workshop_Group 2_29 April 2023)*

There was also an interest to understand how healthcare professionals would support patients and families throughout the procedures and how the patients would be involved in the decision making process:

> *“In which ways will the nurses collaborate with us the patients? While the procedure is ongoing, is it that I will just be receiving orders from the nurse or..?”(Arx Patients Focused workshop_Group 3_29 April 2023)*

### Overall enhanced analysis of perspectives provided by patients and caregivers

The first FGD was conducted two years after the initial camp session, which allowed sufficient time for the research team to support familiarization of advanced therapies among participants, and time for participants to reflect and develop a personal understanding of the implications of the therapies on their own health or that of their family members. As a result, the research team was able to gather in-depth insights on participant perspectives towards advanced therapies for SCD. Table 3 presents a high-level summary of some of the main thematic topics gathered through the qualitative data analysis.

**Table 3.** High-level summary of some of the main thematic topics gathered through the qualitative data analysis.

| Thematic topics | Illustrative quotes from FGDs or IDIs |
| --- | --- |
| **What motivate participants' decisions to accept curative therapies for SCD (HSCT & gene therapies)** | • *“The only thing, I can say here to motivate people is their own personal experiences with the disease.... For instance, some patients can't go more than two months without being hospitalized, which creates a greater need for these treatment” (FGD 1_ Caregiver_speaker 6_ Oct 2023)*<br>• *“I would like to know about the children who were announced to have undergone bone marrow transplantation. How is their progress so far? How are they inspiring us?” (FGD 2_ Caregiver_speaker 7_ Oct 2023)* |
| **How will the costs of treatments be covered?** | • "...the thing that bothers me deep inside is the cost of treatment, that's where the challenge lies. Sometimes I wonder if the treatment is really there, but if we decide to charge, can I afford it? Because there comes a point where you can't even take your child to the hospital or clinic because you don't have the money to pay there. That's the challenge, but basically, I'm happy to see that alternative treatment exists..." (FGD 1_Caregivers_20 July 2023)<br>• "...but what has now discouraged me is about the costs, considering I'm alone, I wonder how I would manage them? The fact that I'm struggling with just affording normal care at this point, worries me. How will I manage to afford these costs for my child to reach this point? That's when I realized it would be impossible..." (FGD 5_Caregivers_19 Oct 2023)<br>• "For my part, I think the government could assist us in a few ways, such as using health insurance to cover half of the treatment costs, with the remaining half being the patient's responsibility. There should be procedures in place for this because, for example, a family member is currently being treated abroad, and we have arranged to pay in monthly installments. This has brought us relief, showing that if such a system exists, it should be utilized. We continued to pay in installments until the insurance was exhausted. Right now, if you don't have money and you're sick, you can't do anything; it means you can't get treated. Many insurance companies are currently refusing coverage. In Tanzania, all our insurance companies refuse to cover certain risks. You might pay a large premium, like 2 million or 3 million shillings, but when you need treatment for something like sickle cell disease, they tell you it's not covered." (IDI 5_patient_13 OCT 2023) |
| **What are the worries and safety concerns related to the treatments?** | • "I'm really scared about what might happen later because of these advanced treatments. What about my kids? Will they also get sick like me because of the genes I pass down to them?" (FGD_Workshop_Patient_29 Apr 2023_Grp 1)<br>• "If I get a transplant from a woman, will it change who I am inside?" (IDI 7_Patient_26 Oct 2023) |
| **Where did patients and families learn about the curative therapies?** | • "For me, I started hearing about advanced treatments after visiting the clinic, where the doctor hinted at it. It's about a year ago. But this year, I've also heard in the media that there are treatments for sickle cell disease, which involve something called stem cell transplantation, that have already been introduced in the country. However, I haven't been able to find out more about whether the patients who underwent the treatment have sickle cell disease, because I've heard that these treatments also treat other conditions." (FGD 1_Patients_27 July 2023_sp 1) |
| **Influence of religious beliefs on the outcome of the treatments?** | • *"I have never sat with religious leaders, but according to the faith, they tell you that illness is something given by Almighty God or that it is how Almighty God created someone, so you cannot change a person's condition or return them to a normal state."*(IDI 11_Caregiver_09 Nov 2023)<br>• *"As a parent, I can say that, from my perspective, religion would not forbid this because it is a form of treatment. Of course we pray to the Almighty God for his help. We will pray for both our child and the doctors, asking God to give them ease and guidance in their work, so they can perform to the best of their abilities under His care. We will turn to our faith to ask for God's support."* (IDI 3_Caregiver_20 Oct 2023) |
| **What supportive network will be needed by families before, during and after the treatments?** | • *"First, let's work together with relatives or friends to prepare your environment because after undergoing treatment, you will be dependent for a certain period. Therefore, until you recover, you need to be under close care. Thus, working with relatives and friends to create a supportive environment can allow one to recover quickly, and some can provide love and affection which can speed up the recovery process and allow one to return to their duties"* (FGD 1_Patients_27 July 2023)<br>• *"You should include family members because this individual is part of a larger family unit, even if the support they offer is limited. It's important to engage adults, particularly close relatives, so they're aware of the situation. Challenges can arise today or in the future that may be too much for one person to handle alone. By involving others, they can offer their perspectives and support. Later on, they may wonder why you tackled everything alone."* (IDI 3_Patient_13 Oct 2023) |

## Discussion

Patients and their families are critical stakeholders in the healthcare system and, thus, incorporating their perspectives into planning activities for novel treatments is arguably compulsory. As such, identifying qualitative research approaches that maximally enable the elicitation of patient insights is important to ensure the success of these types of healthcare programs. The study described in this report combined DE with traditional qualitative research techniques in order to enhance participant engagement and knowledge regarding advanced therapies for SCD, design of study tools, and data acquisition. The results included heightened levels of engagement and successful collection of high quality patient and family perspectives.

Leading healthcare institutions and organizations in Tanzania are helping to herald a new era for SCD care in the country characterized by potential cures achieved through advanced therapies. The first three HSCT procedures performed for SCD in Tanzania, announced by the Ministry of Health in June 2023, constituted a historic milestone. Later that year, the first *ex vivo* gene therapies for SCD were approved by the US FDA. These local and global accomplishments served to animate the SCD community in Tanzania and indeed supported the timeliness and contextual relevance of the current study. Many patients in Tanzania had been exposed to media reports describing curative therapies for SCD, which precipitated their interest in participating and contributing perspectives.

Utilization of efficient engagement approaches is essential to facilitate informed and productive discussions, and is especially important in the context of introducing new interventions in communities where there is a baseline lack of familiarity about them. In pursuit of the overarching goal of using qualitative approaches to explore barriers and facilitators of curative SCD therapies in Tanzania, the research team began its efforts by exploring how best to engage SCD patients and caregivers in discussion about therapies about which they were originally poorly conversant.

The study team had extensive experiences working with SCD communities that provided a foundation for designing the current study, including assessment for how literacy informs understanding of genomic research and the influence of gender in SCD care.^34,35^An important learning from this previous work was that that the quality of research findings depended entirely on the context in which the research was being implemented, and for the topic of the current study we understood the need to firstly conduct facilitated sessions with patients and caregivers to understand their baseline level of knowledge and how best to construct the discussion sessions. Through those experiences, and leveraging the experiences from other research groups utilizing DE approaches to conduct research in other disease areas,^28^ we were able to build a bottom-up approach to further inform the implementation of the qualitative study.

The camps were designed as a hands-on exploratory phase to learn about the extent to which participants would be keen to engage in the discussions around HSCT and gene therapies. Given that a red cell exchange transfusion program was established in Tanzania around the same time, it was a fitting opportunity to extend discussions regarding exchange transfusion to HSCT and gene therapy. In this way, we were able to observe a steady increment of knowledge among participants as discussions shifted from current standards of care to new therapies. The camp sessions were deliberately unstructured to allow a free flow of information (between participants, and between participants and researchers) that enabled researchers to learn how discussions naturally evolved. The workshops that followed the camps were semi-structured to promote discussions that were more focused on the two curative therapies of interest (HSCT and gene therapy).

The experience of the research team was that the DE approach, as adapted for this study, played a key role in familiarizing participants with the new therapies in ways that enabled them to thoughtfully reflect and provide informed analysis based on their own personal experiences living with SCD or caring for a child with SCD. Since all participants that were included in the IDIs or FGDs participated in at least in one DE session, it was straightforward to observe the steady increment of knowledge they gained regarding the advanced therapies. Of note, many participants reported that they learned about curative therapies for SCD only through attending the workshops or the advanced therapy clinic. Participants also provided in-depth perspectives on issues including eligibility for advanced therapies, costs for treatments, and safety. The insights that were offered clearly derived from reflection about participants’ own financial situations and level of access to SCD care. Especially during discussions with patients, worries and concerns regarding safety was a major topic of discussions as compared to FGDs with caregivers.

## Limitations

The duration of the study was longer than originally anticipated because it did require implementation of two methods and analysis for each of the method before moving to the next steps. The focus of the study was mainly patient and caregivers, inclusion of HCPs, could have provided a deeper understanding of the engagement methods. However, despite these limitations, the quality of the outputs was substantially greater.

## Conclusion

It is imperative that patients and their caregivers are provided with knowledge needed to fully understand the implications of novel medical therapies on their own health and, by extension, qualitative research that seeks to understand patient perspectives about advanced therapies must be designed in ways that take into account the potential for variable levels of baseline understanding about the topic. We found that combining DE with traditional qualitative research techniques was highly effective at enhancing the methodology in a study of patient and caregiver perspectives on curative treatments for SCD. The analyses that resulted from this approach are expected to inform the introduction of advanced therapies for SCD in Tanznaia. Furthermore, lessons learned from the methodology can ideally be leveraged to facilitate the design of future qualitative studies in this and similar populations.

## Data Availability

All data produced in the present work are contained in the manuscript

## Ethical Considerations

The study was approved by the Muhimbili University of Health and Allied Sciences Research Ethics Committee. Written consent was sought from the participants for the qualitative study and verbal consent was sought for participation in the deliberative engagement sessions. The study involved research participants who are 18 years and older to ensure that all participants have reached the legal age to consent.

## Acknowledgements

We thank parents and patients who have participated in this research, the Sickle Cell Programme, Department of Hematology and Blood Transfusion at Muhimbili University of Health and Allied Sciences, Aga Khan Hospital, Muhimbili National Hospital, President of Sickle Cell Disease Patients Community of Tanzania, Ms. Arafa Said, Founders of Bone and Blood Foundation, Ms. Neema Mohamed and Ms. Bora Hillary.

